# Restoration of Natural Somatic Sensations to the Amputees: Finding the Right Combination of Neurostimulation Methods

**DOI:** 10.1101/2023.07.16.23292691

**Authors:** Gurgen Soghoyan, Artur R. Biktimirov, Nikita S. Piliugin, Yury Matvienko, Alexander Y. Kaplan, Mikhail Y. Sintsov, Mikhail A. Lebedev

## Abstract

Limb amputation results in such devastating consequences as loss of motor and sensory functions and phantom limb pain (PLP). Here we explored peripheral nerve stimulation (PNS), spinal cord stimulation (SCS) and transcutaneous electrical nerve stimulation (TENS) as the approaches to enable tactile and proprioceptive sensations in the amputees and suppress their PLP. We investigated the efficacy of these approaches in sensory discrimination tasks, namely determining object size and softness using a prosthetic hand. Two transhumeral amputees were implanted for four weeks with stimulating electrodes placed in the medial nerve and epidurally over the spinal cord. Both PNS and SCS induced sensation in different parts of the phantom hand and the corresponding electroencephalographic (EEG) responses. The sensations produced by PNS felt more natural compared to those evoked by SCS. Moreover, neurostimulation-induced sensations were felt as emerging from the prosthetic hand engaged in grasping the objects and recognizing their size. These sensations were further enhanced with dual neurostimulation that enabled appreciation of object softness. The prosthetic sensations became more natural with continued practice. We conclude that the ability to perform complex sensorimotor tasks could be restored to the amputees with an individual-based combination of neurostimulation tools. In addition to restoring sensations, neurostimulation suppresses PLP.

**One Sentence Summary:** The use of peripheral nerve stimulation, transcutaneous electrical nerve stimulation, and spinal cord stimulation improves and enriches prosthetic sensations in amputees by making these sensations more natural and enabling active tasks, such as discriminating object size and softness using a bionic hand.

## INTRODUCTION

The usability of prosthetic devices could be improved with the technology of brain-computer interfaces (BCIs), the systems that connect to the brain to enhance or restore motor functions (1). In BCIs, intentions to perform voluntary movements are decoded from the activity of different areas of the nervous system, which enables control of external effectors even in such complex tasks as handwriting (2). Using electromyographic (EMG) decoders is a common approach to controlling prostheses by the amputees (3).

Even when using sophisticated prosthetic limbs, users still experience difficulties if a prosthesis does not provide sufficient sensory feedback (4). This problem is exacerbated by the presence of phantom limb pain (PLP) experienced by up to 80% of amputees (5, 6). Several approaches have been proposed for improving neuroprosthetic feedback and treating PLP. Peripheral nerve stimulation (PNS) (7) could be used to both implement prosthetic sensations and suppress PLP (8). In addition to PNS, spinal cord stimulation (SCS) is applicable to treat PLP and other types of neuropathic pain (7, 9). PNS and SCS suppress PLP by inhibiting the effects of the pathological discharges generated in the neuromas and affecting the spinal (10) and cortical (11) activity. A preferred neurostimulation system for the amputees would be a bidirectional one where stimulation parameters are set based on PLP-related changes in neural activity (12). In addition to PNS and SCS, such a system could incorporate transcutaneous electrical nerve stimulation (TENS) (13) and targeted muscle reinnervation (TMR) (14).

Notwithstanding the progress made in the development of sensorized prostheses for the amputees (15–17) (8), more research is needed for improving the practicality of such systems. Examples of practical sensorized prostheses include neurostimulation-based solutions for enabling sensations of object size and texture (16, 18). Stability and naturalness of PNS-induced sensations is typically variable in time (19). One approach to improving neurostimulation-based feedback is based on the idea of biomimetics (20, 21) where generation of artificial sensations adheres to the normal somatosensory organization. The embodiment of a prosthesis (22) can be considered as an ultimate criterion of success.

Given that several neurostimulation approaches exist that are capable of generating somatic sensations, it is reasonable to suggest that an individually-adjusted combination of these methods could be particularly effective to enable prosthetic sensations and suppress PLP in the amputees. Accordingly, here we evaluated both the PNS and SCS approaches in two transhumeral amputees. The parameter values of these neurostimulation methods were explored to improve the naturalness of sensations felt in the phantom limb. Furthermore, the patients learned to perform several sensory discrimination tasks with a bionic prosthesis, where PNS and SCS provided sensory inputs. This approach worked well to simultaneously generate proprioceptive and tactile sensations in the tasks where the bionic hand was used to assess object softness. The measurements of cortical evoked response potentials for different types of neurostimulation further clarified how artificial somatic sensations were processed. Overall, these findings improve our understanding of how neurostimulation could provide near-natural multimodal sensations for the prosthetic control while eliminating PLP.

## RESULTS

Two transhumeral amputees, patients S12 and S13, were implanted with the 8-contact electrodes (Directional Lead for the St. Jude Medical Infinity™ DBS System; Abbot; USA) placed in the median nerve and with cylindrical electrodes (Vectris Surescan Trail MRI 1×8 compact 977D260; Medtronic; USA) placed epidurally over the spinal cord at the level of Th6-7 (Fig. 1). In both patients, PNS and SCS evoked sensations in the phantom hand. These sensations spatially overlapped with the zone where PLP was experienced.

**Fig. 1.**
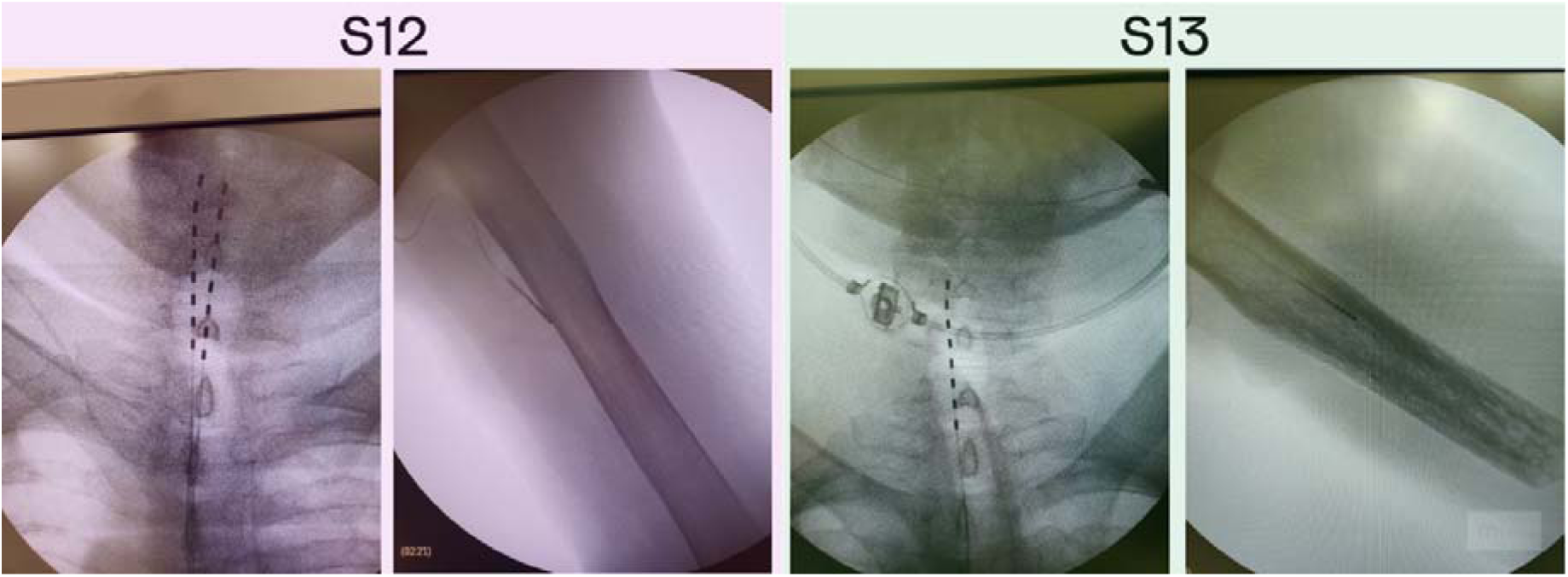
X-ray images of the PNS and SCS electrodes position in patients S12 and S13. (A) Patient S12 was implanted with two electrodes in the area of intumescentia cervicalis of the spinal cord and one electrode in the medial nerve of the left hand. (B) Patient S13 was implanted with two electrodes in the area of intumescentia cervicalis of the spinal cord and one electrode in the medial nerve of the left hand.

### PNS and SCS naturalness assessed with sensory mapping

The sensory mapping of the PNS effects was conducted several times throughout the study. The mapping revealed that for some electrode pairs the location of evoked sensations on the phantom hand remained at the same for as long as 24 days following the implantation surgery.

In patient S12, stimulation with all electrode pairs evoked sensations in the thumb. Additionally, in 19% of cases where 5 different electrodes were engaged, sensations were concentrated in the other fingers (Fig. 2a). A migration of the sensations evoked in the thumb was noticed: they shifted from the base of the thumb to its fingertip and the fingertip of the index finger by experimental day 12. Concurrent with this shift, the patient started to report proprioception-like sensations of the thumb and index finger being flexed during PNS. Additionally, the patient reported naturalistic sensations more frequently. By the third session (day 22), the rate of naturalistic descriptions increased to 33% from the initial 11% (Fig. 2c).

**Fig. 2.**
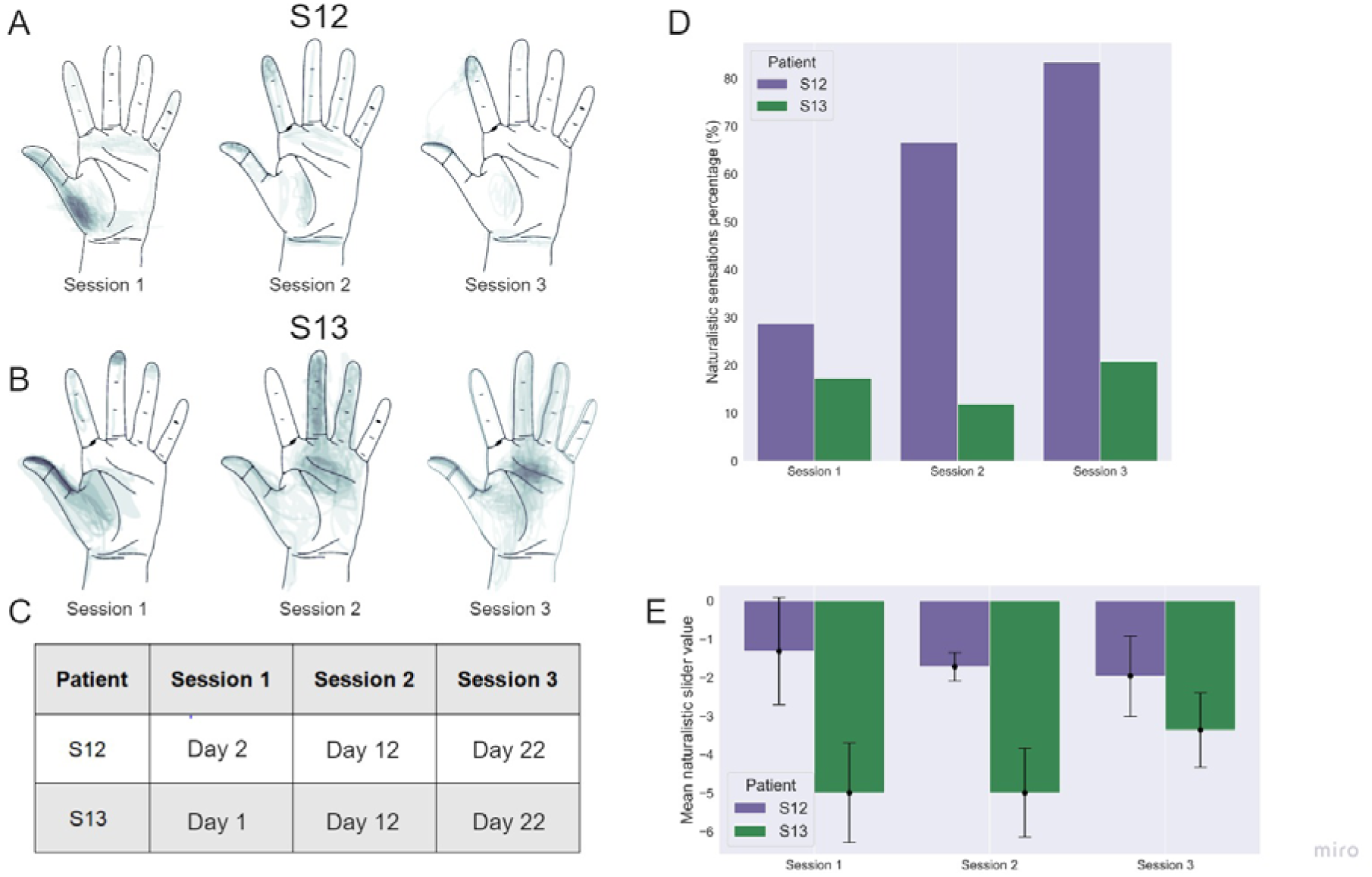
Sensory mapping results. Data for patients S12 and S13 collected in three mapping sessions are shown, conducted with an interval of ∼10 days. (A) and (B): drawings of stimulation projection zones in patient S12 and S13, respectively. Shading corresponds to the density of reported sensation locations. In both patients, the evoked sensations initially concentrated around the thumb but shifted towards the other fingers during the 2nd and the 3rd mapping sessions. (C): - A table showing the dates when the mapping sessions were conducted. Day 1 is the next day after the implantation surgery. (D): Percentage of naturalistic and non-naturalistic sensations by the mapping session. This percentage markedly increased in patient S12 but stayed approximately the same in patient S13. (E): The average value of the analog naturalness slider across sessions. Positive values denote a sensation that was experienced as a natural and familiar to a subject. while, negative values represent odd somatic sensations that the subjects have not experienced during their daily routines

In patient S13, several parameters of stimulation resulted in sensations of touch, pressure and itch (Fig. 2d). A significant increase in sensation naturalness occurred by the last mapping session (Fig. 3e) as compared with the first (Mean diff.=-1.450; p-adj=1.082e-7; Tukey HSD) and the second (Mean diff.=-1.629; p-adj=4.309e-10; Tukey HSD). Additionally, like in patient S12, sensations in the tip of the thumb and the middle of the palm could be evoked with a pair of electrodes (Fig. 3b).

**Fig. 3.**
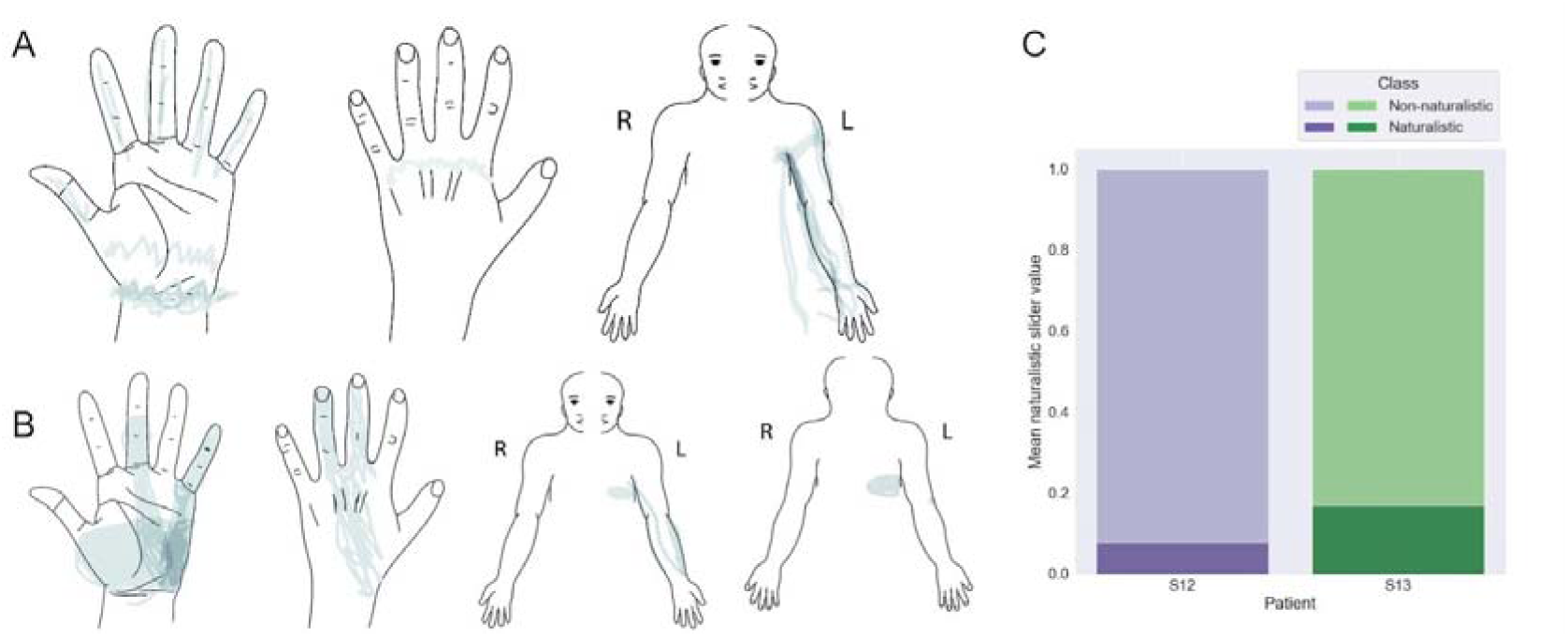
Mapping of sensations evoked by spinal cord stimulation. (A) and (B) maps for patients S12 and S13, respectively. Shading density corresponds to the frequency of reported locations. (C) The proportions of the reported naturalistic and non-naturalistic sensations.

In summary, measurements of several parameters showed that the degree of naturalism of the evoked sensations increased in time in both subjects. Notably, in both participants, we observed a migration of the projection zones from the palm to the fingertips during sessions 2 and 3. This result could be related to neuroplasticity induced by the continued use of neurostimulation including the object-size discrimination tasks.

The comparison of PNS and SCS in terms of naturalness and localization of projections showed that in both patients SCS projected to the phantom hand but also to the arm and trunk. In patient S12 sensations in the phantom palm were evoked in 20% of SCS trials while for PNS each trial was associated with the perceptions in the thumb or pointing finger (Fig. 3a). In patient S13, SCS sensations in the phantom hand were evoked for 36% of the tested electrode configurations, and PNS projected to the phantom hand for 68.8% of the configurations (Fig. 3b).

With respect to sensation naturalness, SCS tended to generate less naturalistic sensations compared to PNS. Only 8% and 17% of the sensations evoked by SCS were marked as natural by patients S12 and S13, respectively (Fig. 3c). Patient S12 rarely reported sensations of hand movements during SCS whereas such sensations were often induced by PNS. In patient S13, SCS and PNS evoked similar sensations.

### Object size detection improves by training day

Both subjects discriminated the size of an object grasped by the prosthetic hand with an accuracy exceeding the chance level of 33% (Fig. 4). On day 11, patient S12 improved the accuracy from 28% to 57%. This improvement had the following steps. During the “Evaluation after training” session, the patient erroneously marked medium objects as large ones. On day 20, the patient did not make this mistake any more without any additional training, so performance accuracy reached 67% which was significantly higher than random performance computed using permutations (Sum of the ranks=0; p_value=7.776e-117; One-tailed Wilcoxon). The entire permutations statistics are presented in Fig. 4. Notably, accuracy for the medium-size objects increased from 6.67% to 23.33%. The Day-20 final score following the training was 73.33% (Fig. 4B). In patient S13, performance accuracy was poorer than in S12 during the first day of the experiment. Before the training, S13 could not match the neurostimulation-evoked sensation with object size and skipped all the trials. Following the training, accuracy reached 34%. On day 20, S13’s accuracy increased to 85% before additional training was conducted. In the control session without stimulation, both subjects were unable to discriminate object size.

**Fig. 4.**
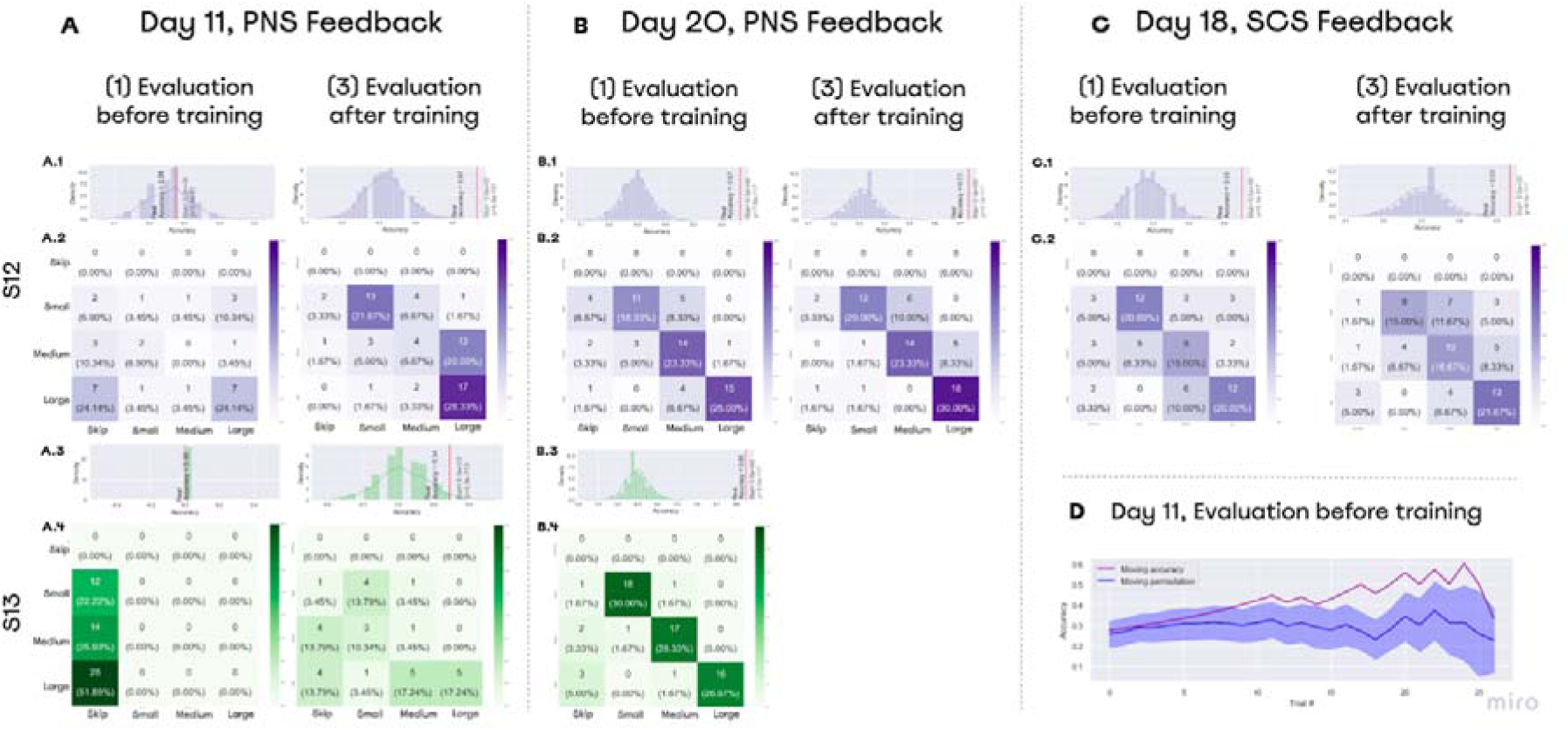
Performance on object size discrimination. (A), (B), (C) - Confusion matrices for object size discrimination. The histograms shown on top of the confusion matrices represent distributions for random accuracy obtained with permutations. Red line represents the real accuracy obtained from the experimental data, and Wilcoxon test statistics are shown. (A) Day 11 data. On that day, both subjects improved their performance following a training session. (B) Day 20 data. Both subjects completed the task with the accuracy exceeding 65% before training was conducted. (D) Day 18 data. Patient S12 discriminated object size with accuracy over 55% using SCS-based feedback. (D) Performance data for day 11. During “Evaluation before training”, patient S12 correctly discriminated object size without any prior training. The floating mean for the accuracy (purple line) is plotted together with the random-performance values computed using permutations (blue line). With 700 permutations, distribution was assessed for performance accuracy on each set of trials. Floating random accuracy is plotted, with transparent shadow representing standard error.

During the “Evaluation before training” session, subjects associated the neurostimulation-evoked sensations to the object size without any previous training. Patient S13 could not discriminate object size without training and skipped all the trials of the evaluation session. Patient S12 attempted to recognize the objects. During the starting trials, S12 was skipping the answers, but then the patient started making correct guesses. Starting from the 10th trial, accuracy exceeded random performance (Fig. 4D). In the SCS sessions, patient S12 recognized objects before training with an accuracy of 55% and after the training with an accuracy of 53% (Fig. 4C).

### Softness detection with combined PNS and TENS

Both subjects were able to perform the softness detection task (Fig. 5). Patient S12 could differentiate objects with an accuracy of 75% before the training using only proprioceptive feedback. After the addition of the second sensory channel, accuracy decreased to 47.5%, which was lower than the chance level of 50% (Fig. 5A). In patient S12, the training procedure was effective and resulted in the accuracy reaching 75% (Fig 5A, 5B). After the task was switched back to the proprioception-only mode, accuracy decreased to 32.5%.

**Fig. 5.**
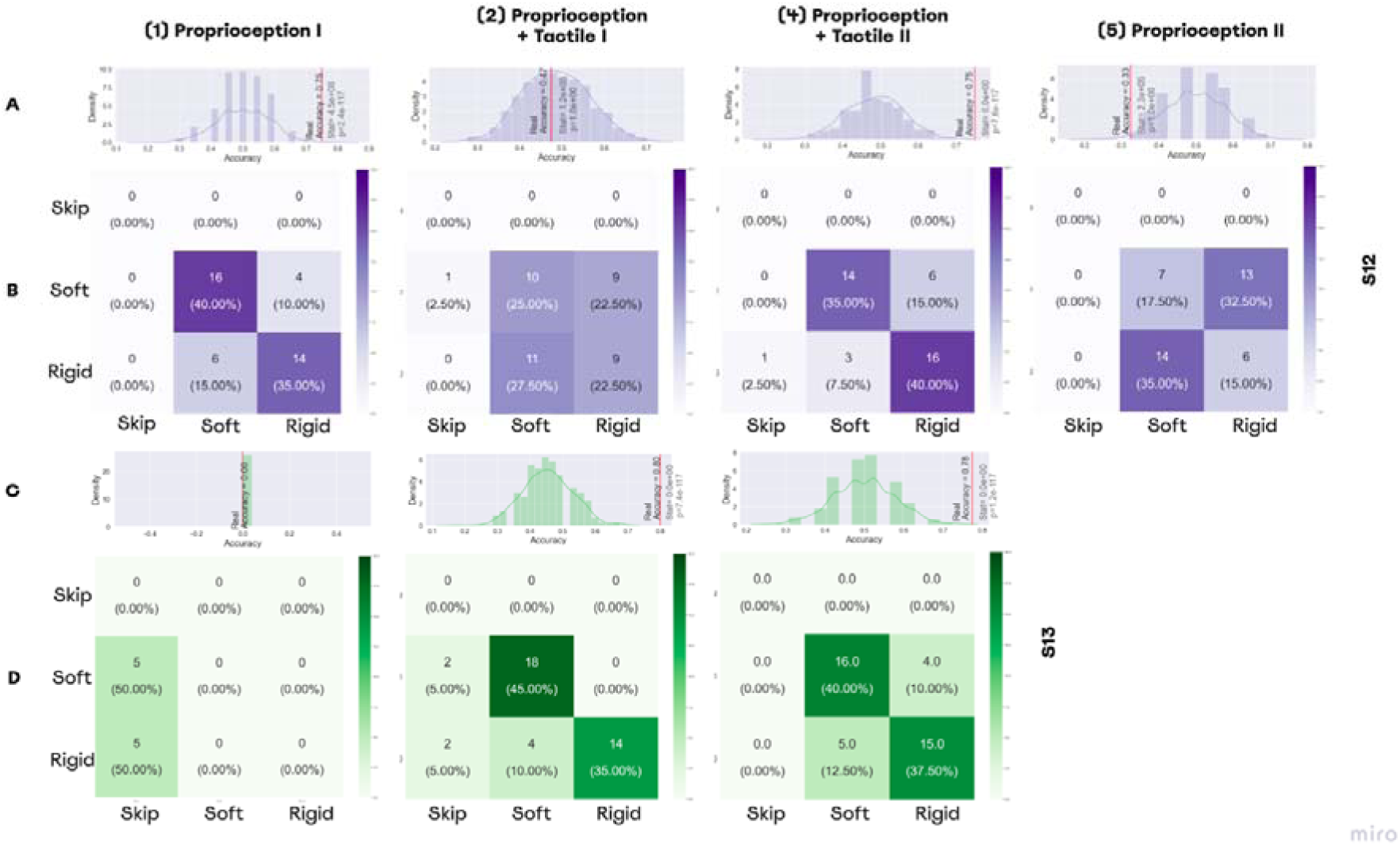
Performance in softness detection task. (A, C) Histograms representing the distributions for random performance accuracy calculated using permutations. The red line indicates accuracy for the actual experimental data. Wilcoxon test results are shown, as well. (A) The distribution for the permutations obtained from the answers of patient S12, (C) The distribution for the permutations for the answers of patient S13. (B, D) The confusion matrices for the sessions of softness detection, (B) The confusion matrices for patient S12, (D) The confusion matrices for patient S12.

For patient S13, the first session was shorter due to technical reasons, and during all trials he was skipping answers without making guesses. By contrast, with an additional PNS feedback that represented the tactile modality, the patient discriminated object size with an accuracy of 80%. Following the training, accuracy was 77.7% for the dual-sensory paradigm (Fig 5C, 5D).

### Embodiment

In patient S12, the embodiment increased from the baseline estimation of −3 to −1 on day 11. The next measurement showed the embodiment decreasing to −6. For the three questions from the control group (Fig. 6), the answers changed dramatically. Similarly, patient S13 changed the answers to five out of six control questions. Though, the “agree” statements increased in number for the target three questions answered by patient S13 too. His embodiment increased from −9 for the baseline estimation to −7 and to −1 for the first and second sessions, respectively. The most prominent change occurred in the answer to “I felt as if the object caused the touch sensations that I was experiencing”, which changed from “Totally disagree” to “Agree”. Notably, both subjects “Agreed” or “Totally Agreed” with the statement that PLP was suppressed during the tests of object size and softness discrimination.

**Fig. 6.**
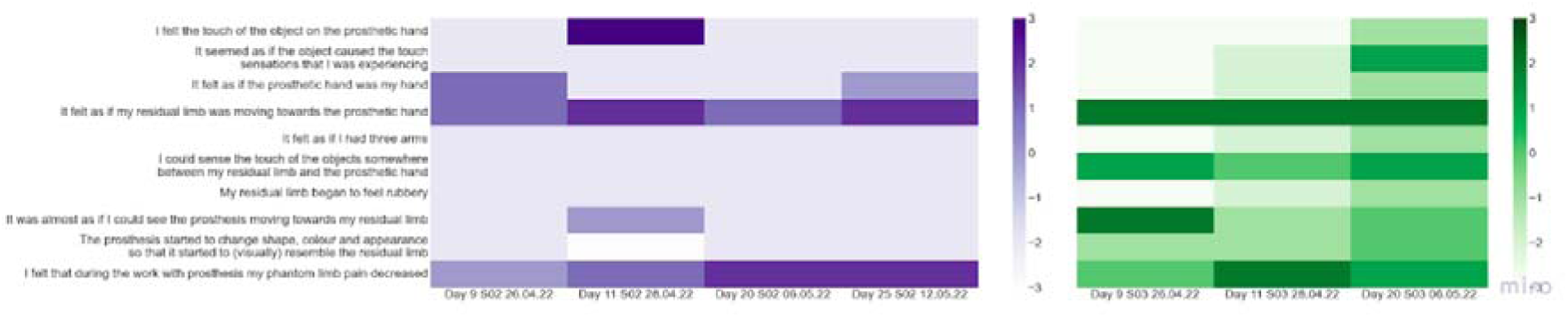
Prosthetic embodiment estimation. S12 and S13 were asked to fill the embodiment questionnaire which estimated if neurostimulation-based feedback had an effect on the prosthetic hand embodiment. These measurements were taken after each test of size and rigidity detection. The baselines estimation of embodiment was measured on day 11. The color of the matrix represents the level of agreement with the statement from −3 (Totally disagree) to 3 (Totally agree).

### ERP lateralization and adaptation to SCS and PNS

In patient S12, stimulation resulted in a clear evoked response potential (ERP). The ERP was stronger for SCS than for PNS (Fig. 7A), and was stronger for the longer stimulation. In patient S13, no clear ERPs were found.

**Fig. 7.**
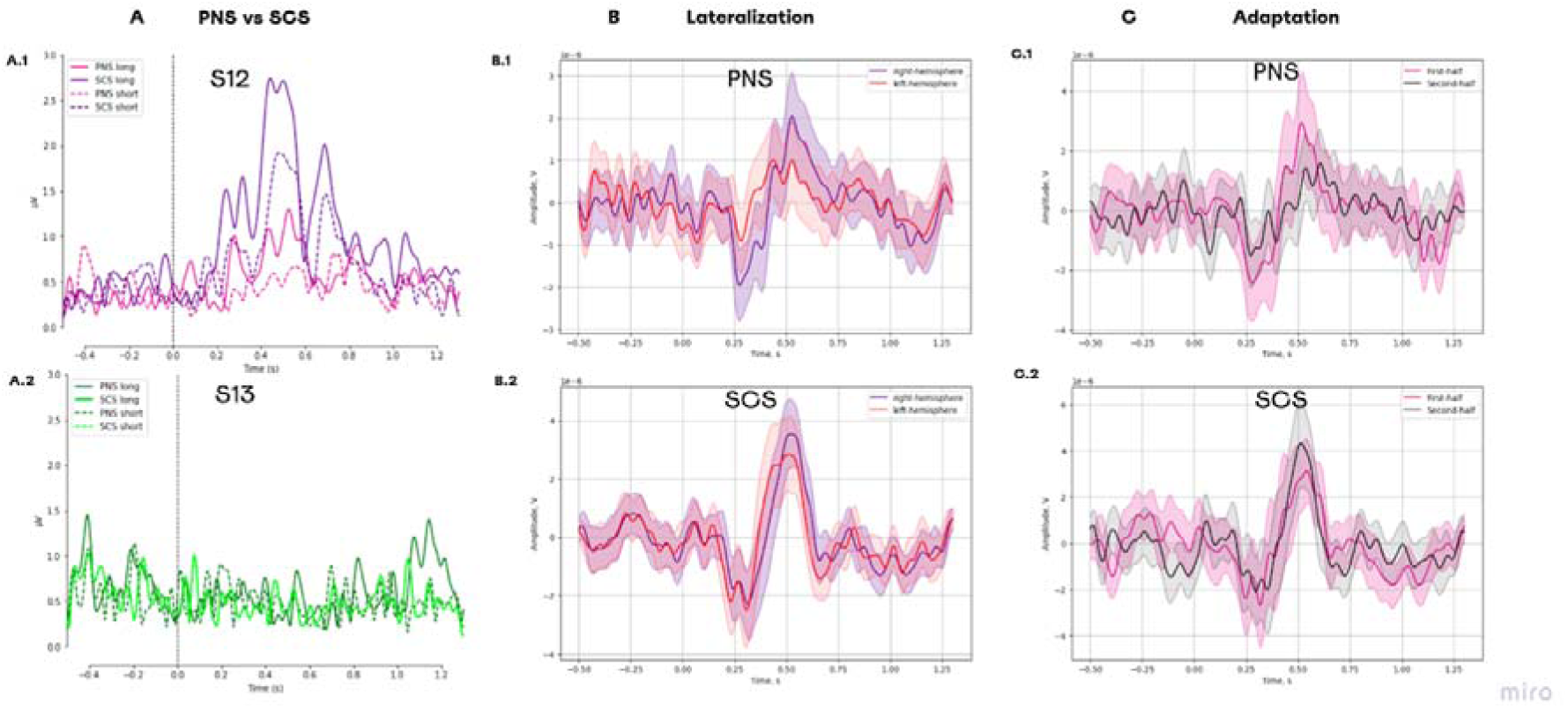
Evoked response potentials for different types of neurostimulation. (A) ERPs for the following conditions: PNS_long, PNS_short, SCS_long, and SCS_short. (A1) Data for patient S12. (A2) Data for patient S13. (B) ERPs in different hemispheres. (B1) The responses to PNS. (B2) The responses to SCS. (C) Change in ERP components over the course of the experimental session. (C1) The responses to PNS splitted into two halves. (C2) The responses to PNS splitted into two halves.

The comparison of long SCS to long PNS revealed a stronger lateralization during PNS for the component N1 (t(98)=-4.605, p = 0.000, paired t-test) and P1 (t(99)=2.892, p= 0.017, paired t-test) (Fig. 7B). For SCS, lateralization was weaker for N1 (t(98)= −0.073, p=0.942, paired t-test) and P1 (t(98)= 2.797, p = 0.017, paired t-test). For PNS, a reduction to PNS was found for the components P1 (t(49)=2.265, p=0.056, paired t-test) which had a higher amplitude for the first half of the trials than for the second half (Fig. 7C). For SCS, no reduction occurred in P1 (t(48)=-1.374, p=0.234, paired t-test).

### Suppression of phantom limb pain

Neurostimulation resulted in PLP suppression in both subjects with the effect average value of 46.00% ± 22.98% (Mean ± St. dev) and 17.20% ± 8.84% in S12 and S13, respectively. Both subjects reported that pain intensity swung during the day without any consistent pattern. In patient S13, PLP further decreased after PNS and SCS were combined on day 13 (Fig. 8). Before this combination was made, PLP suppression was 9.58% on average, and after day 13 it was 24.58% on average.

**Fig. 8.**
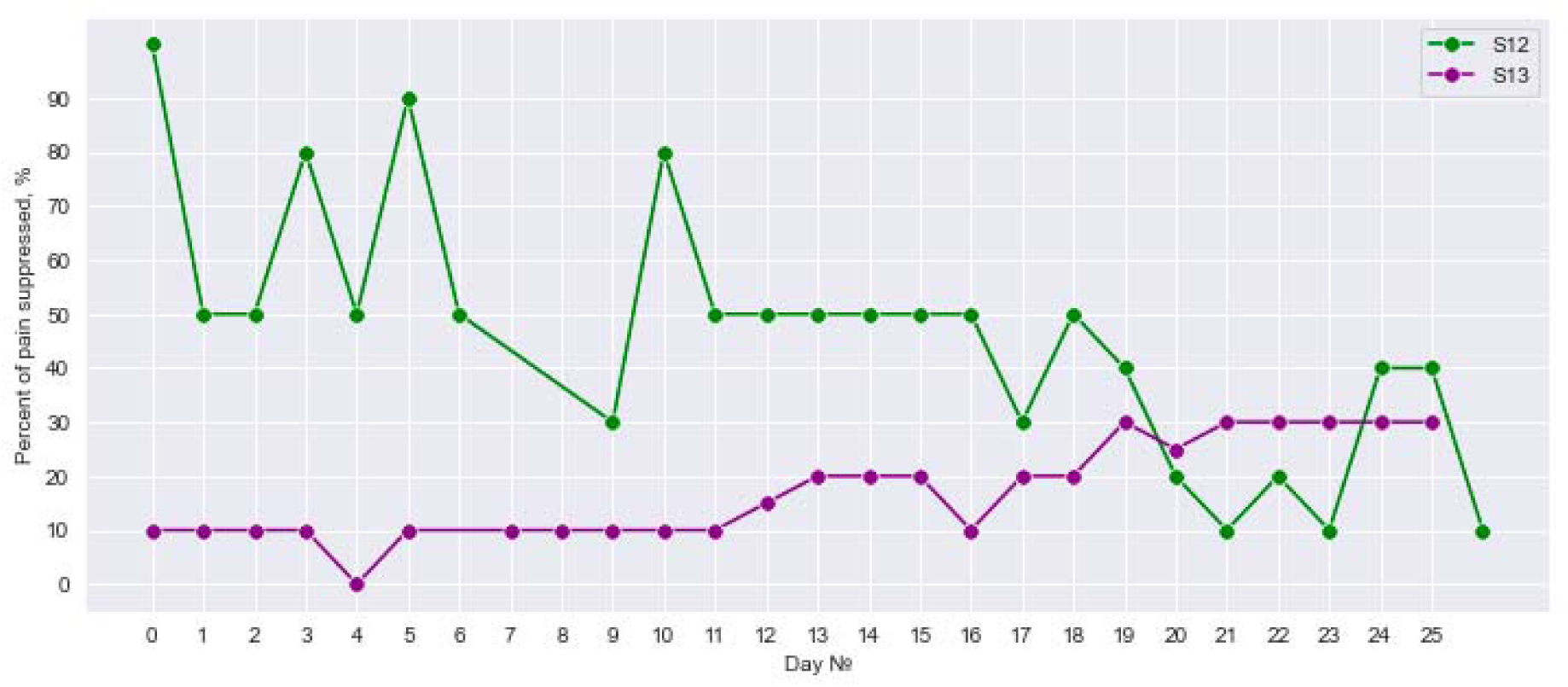
Daily suppression of phantom limb pain. The percent of PLP suppression during 25 days of neurostimulation treatment.

## DISCUSSION

In this study, PNS and SCS evoked somatosensory sensations in the phantom palm of two upper-limb amputees. The naturalness of sensation improved as the patients practiced with neurostimulation, and it tended to be higher for PNS than for SCS. EEG recordings showed that PNS resulted in more lateralized responses as compared to SCS, with a faster adaptation. The neurostimulation-based prosthetic feedback enabled both patients with the capacity to discriminate object size when grasping objects with a bionic hand. Discrimination accuracy improved as the patients practiced on that task. Moreover, by combining PNS mimicking tactile sensations and TENS mimicking proprioception, discrimination of object softness was enabled.

### PNS for neuroprosthetic sensations

Somatosensory substitution systems were previously developed for amputees using vibratory motors and haptics devices attached to the residual limbs (23). Alternatively, PNS-based systems provide feedback of direct phantom limb sensations that enable object size detection and increased prosthetic embodiment (8). Little is known about the progression in time of these discriminative abilities. In the current study, we showed that PNS-based sensations accuracy of sensory discrimination could reach 85% after a prolonged use of the neuroprosthetic system. One of the participants (S12) discriminated the size of three objects without any prior learning on the task. To our knowledge, this is the first demonstration of a PNS-based prosthetic not requiring any prior perceptual training on sensory discrimination tasks. We suggest this is the strongest argument for the possibility to restore naturalistic kinesthetic sensation in the phantom limb using PNS. Such systems are crucial for the upper-limb amputees who need sensory feedback to operate their prostheses (4). During the sensory discrimination tasks, all patients reported that their PLP was suppressed, which makes the developed sensory restoration system beneficial for both sensory substitution and PLP treatment.

SCS is the other invasive-neurostimulation approach to somatosensory feedback (24, 25). We found that with SCS used as sensory feedback, one patient (S12) was able to discriminate object size when three objects were used. This finding supports the approach where SCS generates prosthetic sensations. We added to the previous literature on this usage of SCS a demonstration that PNS, SCS, as same as their combinations could generate different types of prosthetic sensations in the same participant. Moreover, we showed that the experience in sensory discrimination aided by PNS could be transferred to the same task aided by SCS.

### Dual-stream integration

The direct integration of proprioceptive and tactile submodalities of somatosensation is a crucial part of our daily motor behavior. The most that is known about the mechanisms of somatosensory incorporations has been collected in primates studies (26, 27). Lesion of the critical areas (such as Brodmann area 2) result in inability to perform motor coordination tasks (27). Thus, development of neuroprosthetics systems that restore both modalities is key. For example, to identify an object’s softness we require both components since the information about tactile intensity should be associated with the level of the hand’s grasp. Previously, such tasks with amputees were completed only with the systems assembled with the use of TMR and vibromotors (28). We developed the first system that incorporates PNS and TENS for this task. In our study, patient S13 successfully used these two sensory streams to differentiate between soft and rigid objects with the accuracy of 80% even without visual training. Notably, he was unable to complete this task using only TENS that mimicked the proprioceptive domain. Surprisingly, S12 was able to differentiate between two objects with different softness using only PNS feedback that mimicked the aperture level. We observed that S12 adapted to PNS as a signal for aperture encoding during the object size detection sessions. Since the soft object diameter decreased by only 55mm and the rigid object diameter by 60mm, we conclude that the patient learned to detect even slight changes in PNS amplitude which could be beneficial for many manual tasks. Alternatively, when both types of feedback were combined, his performance decreased to 47.5%, but after visual training it again increased to 75%. This rapid adaptation to the new types of feedback can be explained by effective perceptual learning (29).

### Naturalness

The naturalness of evoked somatosensory sensations as a phenomenon in the somatosensory restoration has been mainly assessed with questionnaires (19, 25, 30). To achieve a higher naturalness, in some studies the stimulation current is modeled to mimic the normal sensory nerve action potential (SNAP) (21). Such biomimetic approaches seem to provide more natural feedback and allow a higher performance in motor tasks with a prosthesis (31).

Another approach to enhance naturalness using PNS stimulation is the development of specific electrode configurations that provide focal stimulation of distinct nerve fibers (32–34). Yet, even with the development of transverse intrafascicular multichannel electrodes (TIME) it is still unknown how to localize the electrode, so it causes activation of nerve fibers of expected specificity (35). The methods of immunofluorescent staining allow to differentiate afferent and efferent fascicles (36), though it cannot be applied for the field of neuroprosthetics.

By contrast, in the behavioral data we observed that naturalness increased in time without specific affection. In the only case study, (19) reported that the sensation of naturalness was increasing during daily use of PNS-based prosthesis during the first month of use. We observed a similar trend of growing naturalness for the sensory mapping sessions for S12 and S13. This pattern of increasing naturalness resembles the effects of cochlear implantation (37). Notably, patients who use cochlear implants experience mechanical and high-pitched sounds during the first 3-6 months of use. Thus, in the first six months rehabilitation implies active adaptation to the new way of sound. Additionally, during this period, their devices are being adjusted by a team of clinicians. Such changes in natural perception may be explained by the mechanisms of sensory normalization and sensory adaptation which exist in the somatosensory domain too (38). Some short-term adaptation to PNS stimulation was indicated in this study using EEG ERP, but additional long-term changes are in high interest for the following studies.

Since our patients were implanted with both PNS and SCS electrodes, we are the first to show the difference in sensations induced by these types of neurostimulation for the same participants. S12 reported the sensations felt as more natural in PNS sensory mapping. And this might have happened because medical specialists applied PNS as the main neuromodulation technique for his PLP suppression which wasn’t the case for S13. To summarize, we suppose that sensation naturalness can be obtained by: (1) appropriate spatial localization of electrodes, (2) biomimetic current characteristics and (3) rehabilitation that induce sensory adaptation.

### Embodiment

One of the key benefits of PNS and SCS based feedback in neuroprosthetics is an increased sense of embodiment that was reported in many case studies (16, 39). The sense of embodiment is also important since the increasing number of wearable devices and attempts to full immersion into virtual reality (40). The complex phenomenon of embodiment seems to have an ambiguous definition (22). From one perspective, it is defined as the process by which a foreign object becomes integrated into the existing neural infrastructure that supports the body. On the other hand, it is described as “subjective experiential correlate” that implies the accumulation of a sense of ownership and sense of agency (22).

In our research, we estimated it using the questionnaire from (41), which is a common approach for estimation of prosthesis embodiment. It was originally developed from a rubber hand illusion (RHI) questionnaire, and it contains a set of target and control questions. Surprisingly, we observed that in both subjects agreement increased mainly in the set of control questions making the result of the questionnaire controversial. We explain this by inexperience in prosthesis usage in our participants during the first measurement of embodiment. None of them used prosthesis before our study, making the baseline estimation of embodiment inaccurate. Though, some critics to the control questions of RHI questionnaire were already mentioned, especially for studies with amputees (42). To avoid such issues, we highly suggest switching to an implicit way of embodiment estimation, using such phenomenons as sensory attenuation or cross-modal congruency (22). Ultimately, the sense of embodiment might be associated with naturalness of sensation, but additional research is needed in this topic with the use of objective methods to prosthesis embodiment estimation.

### EEG

The direct stimulation of peripheral nerves is a well-known method for the investigation of somatosensory ERP (43, 44). It was not previously used for the comparison of effects of PNS and SCS. We observed that PNS has a more lateralized response in comparison with SCS. This effect is in high agreement with the behavioral data of lateralized responses that were collected during PNS sensory mapping. The reported decrease in P1 component during the recording for PNS stimulation seems to be the result of somatosensory adaptation (45). We did not collect data about the stimulation perception during EEG recording to correlate the perceived intensity with the component’s amplitude, but such direct association was shown in previous studies (46). These electrophysiological markers of stimulation could improve sensory mapping procedures that will be employed in the next iteration of research. Also, the objective representation of stimulation could be used for a closed-loop neurostimulation that will adjust stimulation amplitude to cause the required level of perceived sensation. Previously, we suggested using EEG biomarkers to adjust stimulation for the treatment of PLP (12).

### Pain suppression

The use of PNS as a tool for PLP suppression is still in need of additional validation (9). We demonstrated the efficiency of such stimulation in our previous study (8) and we added the case of S12 here. Remarkably, in S13 an additional decrease in PLP level was observed when simultaneous PNS and SCS were included in his treatment program. Classically neuromodulation for neuropathic pain is treated within one paradigm of stimulation. Thus, a combination of two types of neurostimulation could improve the treatment.

## MATERIALS AND METHODS

### Surgery and patients

Two amputees participated in the study, and both suffered from PLP. S12 and S13 had transhumeral amputation on the left side. The patient IDs are not known to anyone outside the research group. More details about these cases can be found in Supplementary Table S1. The study was approved by the Ethical Committee of the Far East Federal University (FEFU) Biomedicine school (Protocol #4; April 16, 2021). Each patient signed the informed consent form prior to participating in the experiments. The study is registered as a clinical trial on platform https://clinicaltrials.gov/ #NCT05650931.

Implantation surgeries were performed at the Medical Сenter of FEFU. Eight-contact electrodes (Directional Lead for the St. Jude Medical Infinity™ DBS System; Abbot; USA) were implanted in the median nerve in all three patients under endotracheal anesthesia. Additionally, in both patients electrodes were implanted in the area of intumescentia cervicalis of the spinal cord (Fig. 1). SCS cylindrical electrodes (Vectris Surescan Trail MRI 1×8 compact 977D260; Medtronic; USA) were implanted under local anesthesia and X-ray control.

To implant the PNS electrodes, the epineurium was cut under a surgical microscope. The electrodes were placed in the space between the nerve fascicles. For spinal electrodes implantation, a puncture of the posterior epidural space was performed at the level of Th6-7 under local anesthesia. In patient S12, the SCS electrode migrated to the midline on the 3rd day after the surgery. To correct this issue, an additional electrode was implanted over the spinal cord.

In all patients, sensations evoked by stimulation covered the zone where PLP was experienced. Stimulation paradigms were individually adjusted in each patient to maximize the PLP treatment effect. As a result of these adjustments, patient S12 was treated with PNS, patient S13 received SCS treatment. The stimulation parameters were selected individually and made up 40-100 Hz of frequency and 100-1000μs of pulse width. Patients used diaries to daily mark the level of PLP suppression using a visual analogue scale.

### Sensory mapping

#### PNS

PNS evoked sensations in the phantom hands of both participants. Sensory mapping was the key procedure in which we revealed connections of stimulation parameters with evoked sensations. During this experiment we switched between the set of electrode pairs varying amplitude of pulses in accordance with the predefined algorithm. The mapping was performed three times during three consecutive days. Stimulation frequency was 50 Hz on day 1 and 100 Hz on days 2 and 3. The frequency was raised to 100 Hz because both participants reported more natural sensations for this type of stimulation.

In a trial of sensory mapping an electrode pair was selected, with stimulation parameters set to the initial values (bipolar stimulation with pulse width of 100 μs and frequency of 50 or 100 Hz, depending on the day of testing). Stimulation amplitude was then gradually raised from 0 with 0.1-mA steps. For each of these steps, participants were asked to report sensation intensity on a 0 to 10 scale where 0 corresponded to an absence of perception and 10 corresponded to an uncomfortably intense sensation. The tests with an increasing stimulation amplitude continued until the score of 10 was reached. For the reported stimulation intensity equal to 5, patients were asked to fill a PerceptMapper questionnaire (47). where they marked the perception location on a drawing of a hand shown on a computer display and described the sensation in terms of painfulness and naturalness (8). When the application form was filled, we switched to the next trial where another electrode pair was selected and tested.

The participants used sliders to report the naturalness of the stimulation-evoked sensation naturalness, and the terms suggested in the sensation descriptors tab of the questionnaire to describe these sensations. Two classes of sensations were reported by the patients: naturalistic and non-naturalistic. Sensations of pressure, touch, prick, shock, urge to move and itch were categorized as naturalistic. Sensations of electric shock, pulsing, vibration and flutter were categorized as non-naturalistic. Statistical distributions were calculated for the counts of different kinds of reported sensations. Normalization by the number of trials per day was applied. This analysis was used to assess the efficacy of PNS for evoking natural sensations. To measure the changes in evoked sensations that occurred over the course of these experiments, sessions of sensory mapping were conducted in the beginning, in the middle and at the end of the study. In these sessions, we determined referred sensation sites and the descriptions of sensations and their intensity.

To compute the density of neurostimulation projection sites on the phantom hand, averaging was conducted for the hand images where the patients marked their phantom sensations. The image size was of 1080×1080 pixels, and pixel intensity was in the range from 0 to 255. Since the patients did not mark the sensation zones precisely, the intensity represented the percentage of presence for each pixel. An average image was calculated for each mapping session.

#### SCS

Since both patients were implanted with the electrodes for spinal cord stimulation, we could compare the effects of SCS and PNS. SCS mapping was arranged the same way as the mapping using PNS, including hand images for mapping, reports of the felt intensity of stimulation with sliders, and sensation descriptors tab. SCS mappings were conducted once for each patient on the 3rd and 8th days of testing in S12 and S13, respectively.

### Object size detection

#### Experimental design

In the task that required discriminating an object size using PNS feedback, rigid cylinders made of PLA plastic of different diameters were grasped by a prosthetic hand. Simultaneously, the signals from the prosthetic sensor of aperture were converted into PNS patterns. The cylinders were shown to the patients before the experiments. They came in three sizes: small (20mm in diameter), medium (40mm) and large (60mm). The patients controlled the prosthesis grasping using an external controller with two buttons: “open” and “close”. When a patient initiated the grasping movement, the change of prosthesis aperture led to increased neurostimulation amplitude respectively (See section Stimulation settings for sensorimotor tests for details). We call the sensations they experienced artificial proprioception because they mimicked sensing of hand configuration. In this artificial proprioception, measurements from prosthetic finger encoders provided the information about the prosthesis aperture to a PC via a serial connection with a galvanic isolation.

The object-grasping sessions were conducted on postsurgery days 11 and 20 (Supplementary Fig. S2A). Each session consisted of three parts: (1) evaluation before training, (2) training, and (3) evaluation after training. During the evaluation sessions (parts 1 and 3 of the session), subjects wore a blindfold and noise-isolating headphones (3m Peltor) (Supplementary Fig. S2B). Each patient was instructed that an object would be grasped by the prosthetic hand and his task would be to determine whether the object was small, medium or large. Each trial started with PNS amplitude being lower than the sensory threshold. A new cylinder was placed by an experimenter in front of the prosthetic hand in randomized order, which took about 3 s. Then, the experimenter gently taped the patient’s intact arm with a pen to indicate the beginning of a trial. Total set of trials was 60 per each evaluation step.

Alternatively, during the training period (part 2 of the session), participants had a full vision of the prosthetic hand and the cylinders. They could also hear the sound of the prosthetic-hand motor and they were allowed to freely interact with the objects.

Notably, during evaluation before training (part 1) the subjects did not have any prior knowledge in this task; however they experienced the dynamic range of evoked sensations during the initial amplitude tuning step. Thus, it was tested if subjects can by associating the level of PNS magnitude to the level of prosthetic aperture guess the object’s size. We hypothesized that the stimulation could be naturally matched with the natural kinesthetic sensation in the palm, and a subject could recognize an object even without any prior training.

#### Statistical analysis

For each session, accuracy metrics and confusion matrices of object size prediction were computed. Permutation tests were used to assess statistical significance of prediction accuracy. Namely, the subjects’ answers were randomly shuffled 700 times to compute a statistical distribution of random accuracy. Next, the real experimental accuracy was compared with the permutation distribution to obtain the p-value. We used the Wilcoxon test from Scipy library to compare the mean accuracy from the random permutations with an actual value of accuracy observed in a session of object size recognition.

### Softness detection

#### Experimental design

In the softness detection test, we determined how well patients could use a combination of PNS and TENS to discriminate objects of different softness. A soft object was assembled by wrapping a 20-mm rigid core made of PLA plastic with a soft multilayered shell made of foamed polyethylene sheets of 2-mm thickness (Supplementary Fig. S3A). For the rigid object, the inner core of about 58 mm in diameter was covered by a single layer of a foamed polyethylene sheet. Both objects in non-compressed state were approximately 60 mm in outer diameter.

The neurostimulation was constructed to mimic two somatosensory modalities. For S13 the TENS component served to mimic proprioception, which provided a patient with the information about the aperture of the prosthetic hand. The PNS component mimicked tactile sensations from the fingertips, and the appropriate signals were derived from the pressure sensors of the prosthetic fingers. For S12 we used an opposite encoding, particularly PNS mimicked proprioception and TENS mimicked tactile submodality. For both TENS and PNS, amplitude modulation was used to convey the signal to the patient (for additional details look at section Stimulation settings for sensorimotor tests). We decided to link PNS to the proprioceptive domain for S12 and tactile domain for S13, because during the sensory mapping procedure they tended to associate the PNS with the respective sensations.

The sequence of task events was similar to the object size detection task. An experimenter placed an object in front of the prosthetic hand in a randomized order and then gently taped the patient’s arm to signal the trial start. The patient next pressed a button, which started the prosthetic grasping and neurostimulation.

The experimental session consisted of four sessions of object softness recognition with a training procedure in between (Supplementary Fig. S3B). We call the first part (1) “Proprioception I” because only the proprioceptive mode was turned on, and a subject needed to differentiate objects using only PNS (for S12) or TENS (for S13) feedback. In the second part (2) “Proprioception plus Tactile I” we turned on both stimulation allowing the patient to have both sensory streams during recognition of object rigidity. A patient was wearing a mask and soundproof headphones like in the Object size detection task. Prior to these sessions, the subject did not have any experience in this task, so we consider the results of these two sessions as a pure performance without any perceptual learning.

Then, the subject was free of headphones and mask to have a (3) training session. During this period, he was allowed to freely manipulate objects having both TENS and PNS encoded both somatosensory sensations. Afterwards, again a patient wore a mask, headphones and completed the session of (4) “Proprioception plus Tactile II” when both TENS and PNS amplitude modulation guided the subject during object softness recognition. Finally, one of the simulations was turned off and with remaining PNS (for S12) or TENS (for S13) that encoded proprioceptive domain intensity he completed the session (5) “Proprioception II”. Total set of trials for each step amounted to 40.

#### Statistical analysis

Similarly to Object size detection, accuracy metrics and confusion matrices of recognition were computed. Permutation tests were used to assess statistical significance of recognition accuracy as same as in Object size detection. Then, the Wilcoxon test from Scipy library was applied to compare the mean accuracy from the random permutations with an actual value of accuracy observed in a session of object size recognition.

### Stimulation settings for sensory discrimination tests

For each test of object recognition, stimulation parameters were selected based on the preceding sensory mapping. An electrode pair was used that proprioceptive sensation in the phantom hand. The range of stimulation amplitude was chosen to be comfortable to the subject. This range corresponded to psychometric-scale values from 1-2 (barely perceivable) to 7-8 (massive yet comfortable perception). The readings from prosthetic finger encoders provided a measure of the prosthetic-hand aperture, which changed from 0% (fully closed) to 100% (fully opened). This signal was converted into the amplitude of PNS or SCS with respect to the individual sensory maps. In the Object size test, the proprioceptive information controlled level of PNS in sessions with S12 and S13, and SCS for one experimental session with S12.

In the Softness detection, finger encoders were combined with the pressure sensors (Optical Tensometers by Motorica LLC) located on the prosthetic fingertips. The sensors’ readings were scaled according to the task so that 0% corresponded to a free open prosthetic arm, while 100% — to the highest pressure measured during grasping of test objects. The signals from the pressure sensors were converted into the TENS (for S12) or PNS (for S13) amplitude within the range that was comfortable for the patients. The signal from the pressure sensors were converted into the stimulation amplitude with a linear transfer function where stimulation amplitude did not exceed the psychometric values of 7-8 (Supplementary Fig. S4). The corresponding processing was done using a laptop computer and NimEclipse simulator. Programmatically, aperture was sampled at 30Hz and pressure readings were sampled at 100 Hz whereas the PNS and TENS amplitudes were updated variably at 10-30 Hz depending on the stimulator parameter update latency. In addition, due to the hardware requirements of Medtronic NimEclipse Intraoperation Monitor, the highest frequency for triggering the stimulation trains was 4.6 Hz for two-channel stimulation. Accordingly, this frequency was used for simultaneous PNS and TENS usage. The stimulation was arranged as trains composed of 10 pulses with a width of 100 µs presented at 100 Hz. The trains were presented at 4.6 Hz.

### EEG recordings

#### Experimental design

EEG recordings were conducted using the standard 10-20 montage system with 32 channels. An NVX-136 amplifier (Medical computer systems) was used. The ground electrode was located in the forehead. The mean of A1 and A2 channels was subtracted as a reference.

A patient was comfortably seated in a chair while neurostimulation was delivered. Electrical stimuli with constant frequency (100Hz) and pulse width (900mcs) were provided using peripheral nerve stimulation (PNS) and spinal cord stimulation (SCS). The amplitude was selected individually according to the subject report. Mainly, we replicated the procedure described in the Sensory mapping section and selected amplitudes that caused sensation at the level 5-7 from psychometric scale.

The recording was held in two sessions that differed from each other with the duration of stimulus. For S13 stimuli were 1s (Long) and 500ms (Short) while for S12 they were 1s (Long) and 300ms (Short). Each stimulus appeared with the subsequent synchro impulse that was transmitted via LSL protocol from the experimental software to the Neorec Software.

During the experimental session, each stimulus was presented 100 times with the variable interstimulus interval (3.6 - 4.4 seconds). A Python-based software was used to control experimental sequences.

#### Processing of EEG

The data analysis was performed using MNE Python (48). Data was notch filtered at 50 Hz and bandpass filtered (1-40Hz). Then, ICA was applied, with the use of the ALICE toolbox (49) to remove the artifactual components of eye movements. Noisy channels were dropped from the recording. Then, epochs for each condition were extracted using threshold-based (150μV) removal of contaminated epochs. All epochs were splitted into four conditions PNS_long, PNS_short, SCS_long and SCS_short. The evoked response potential (ERP) for each condition was computed by averaging over remaining epochs. We estimated the components N1 and, as an average value in the window of 30ms of negative peak in ERP and P1 peak in the window of around positive peak.

#### Statistical analysis

For the condition PNS_long and SCS_long, we calculated the value of lateralization. We took an average activity in the region of interest (ROI) area of the somatosensory cortex: electrodes ‘C4’, ‘CP2’, ‘CP6’ of the right hemisphere and ‘C3’, ‘CP1’, ‘CP5’ of the left hemisphere. We calculated the amplitudes of components N1 and P1 between the two hemispheres for PNS and SCS separately.

Additionally, we examined the change in the ERP components over the course of the experimental session. The data for conditions PNS_long and SCS_long was splitted into two halves. Next, we compared the first half to the second to obtain a measure of response change over time. All statistical comparisons were conducted using the paired t-test programmed in Python Scipy. Then, all statistics were adjusted using FDR correction for multiple comparisons.

### Embodiment

To test whether neurostimulation-based feedback had an effect on the prosthetic hand embodiment, S12 and S13 were asked to fill the appropriate questionnaire (41). The questions were translated to Russian because the patients were native Russian speakers. The questionnaire was completed four times:

(1) On day 9 (baseline estimation) when the patients used the prosthetic hand for the first time without sensory feedback in S12 and S13
(2) On day 11, that is after the first session of Object size detection in S12 and S13.
(3) On day 20, that is after the second session of Object size detection in S12, and after the second session of Object size detection and the session of Softness detection for S13.
(4) On day 25, that is after the session of Softness detection in S12 only.

The survey quantified a participant’s agreement with 9 statements: 3 statements of predicted phenomena (like “I felt the touch of the investigator on the prosthetic hand”) and 6 control statements (like “I could sense the touch of the investigator somewhere between my residual limb and the prosthetic hand”). We added one more statement: “I felt that during the work with prosthesis my phantom limb pain decreased” to estimate the state of phantom limb pain during the active tasks. The subject estimated the level of agreement with the statements on the scale from −3 (totally disagree) to 3 (totally agree).

### List of Supplementary Materials

Present a list of the Supplementary Materials in the following format.

Materials and Methods

Table S1

Fig. S2

Fig. S3

Fig. S4

## Data Availability

All data are available in the main text or the supplementary materials.

https://github.com/gurasog/Restoration_of_Natural_Somatic_Data

## Author contributions

Conceptualization: GS, AB, MS, ML

Methodology: GS, AB, MS, ML

Investigation: GS, AB, NP, MS

Visualization: GS, NP, MS

Funding acquisition: AK, ML

Project administration: GS, AB, YM, MS, ML

Supervision: ML

Writing – original draft: GS, NP, AB, MS

Writing – review & editing: GS, ML

## Competing interests

AB, YM, and MS are employees of Motorica LLC, other authors declare that they have no known competing financial interests or personal relationships that could have appeared to influence the work reported in this paper.

*Motorica LLC is a private company, developing and producing functional prosthetics of upper limbs.

## Data and materials availability

All data are available in the main text or the supplementary materials or via link https://github.com/gurasog/Restoration_of_Natural_Somatic_Data

