## Supplemental File 1. for "Restoration of Natural Somatic Sensations to the Amputees: Finding the Right Combination of Neurostimulation Methods"

| Subject ID | S12 | S13 |
| --- | --- | --- |
| Sex | M | M |
| Amputation type | Unilateral left-side transhumeral | Unilateral left-side transhumeral |
| Date of amputation | July 2020 | September 2021 |
| VAS scale | 7-8 | 7-8 |
| DN4 | 3 | 8 |
| Pain detect | 19 | 25 |
| HADS1 | 11 | 2 |
| HADS2 | 5 | 5 |
| SF36 Physical | 40.01394 | 38.34537 |
| SF36 Mental | 43.35149 | 37.34705 |

**Supplementary table S1. Extended information about patients**

Both subjects completed five clinical questionnaires prior to the surgery. Visual-analogue scale (VAS), DN4, and Pain detect were collected to estimate the level of neuropathic pain. HADS and SF36 were collected to estimate patients’ mental condition and quality of life.
