## Supplemental File 2. for "Restoration of Natural Somatic Sensations to the Amputees: Finding the Right Combination of Neurostimulation Methods"

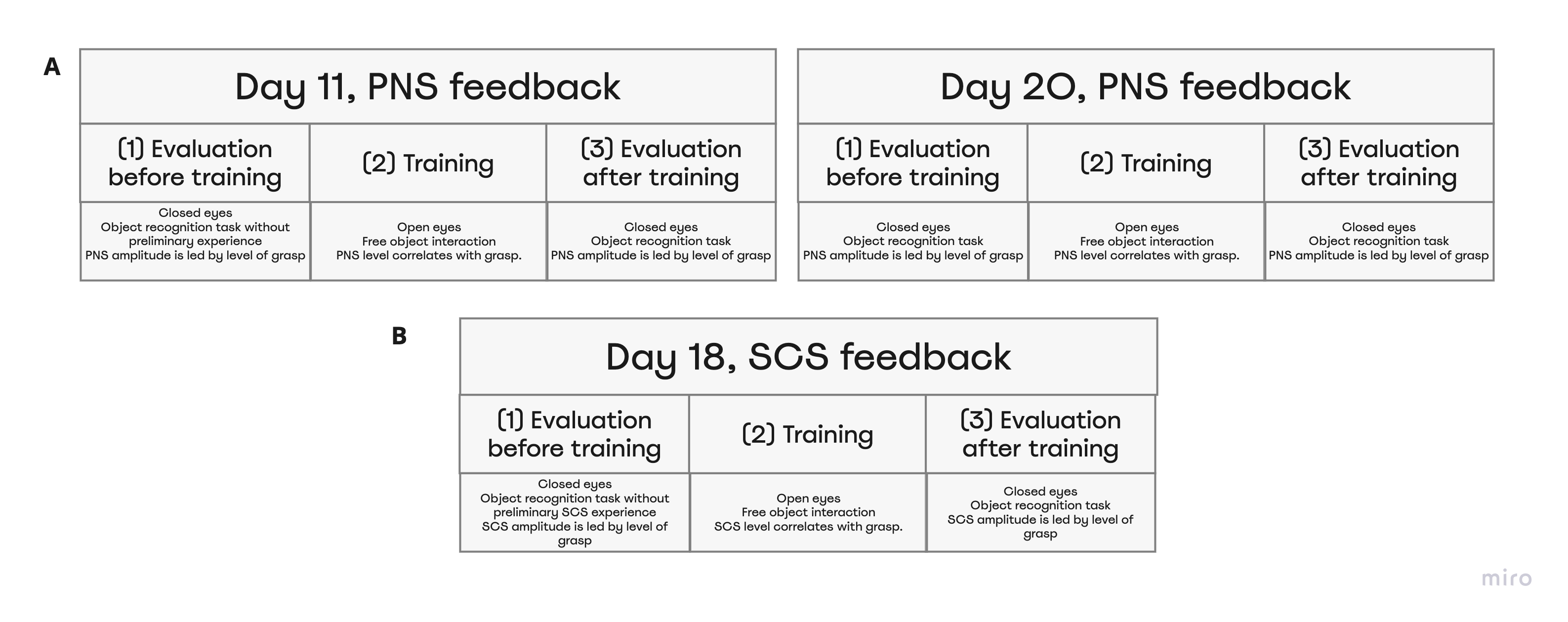

**Supplementary figure 2. Object size detection experimental design and chronology.**

Object size detection was conducted thrice on day 11, day 20, and day 18. Each time experiment was held in three stages: Evaluation before training, Training, and Evaluation after training. Before training and after training subjects differentiated objects of different sizes using neurostimulation feedback that caused a sense of phantom hand grasping. **(A)** - During day 11 and day 20 S12 and S13 completed the task using PNS. **(B)** - During day 18 S12 completed the task using feedback from SCS.
