## Supplemental File 3. for "Restoration of Natural Somatic Sensations to the Amputees: Finding the Right Combination of Neurostimulation Methods"

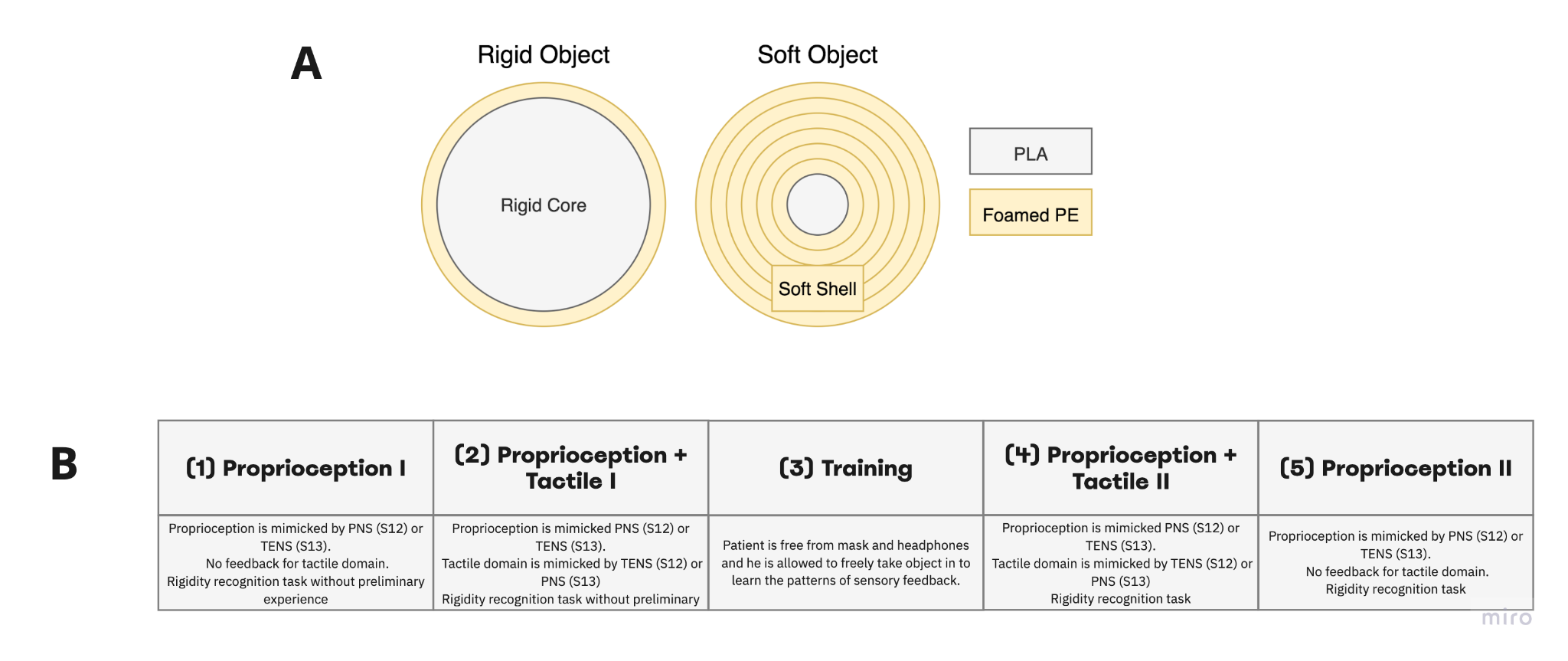

**Supplementary figure S3. Softness detection experimental design.**

**(A)** Participants needed to differentiate between rigid and soft objects. The soft object was assembled by wrapping a 20-mm rigid core with a soft foamed polyethylene of 2-mm thickness. For the rigid object, the inner core diameter made up about 58-mm. Both objects in non-compressed state were approximately 60 mm in outer diameter. **(B)** Experiment was conducted in four stages: Proprioception I, Proprioception plus Tactile I, Proprioception plus Tactile II, Proprioception II. During the experiment, the subjects differentiated object rigidity using PNS (S12) or TENS (S13) stimulation only in sessions of Proprioception I and Proprioception. In sessions of Proprioception plus Tactile I and Proprioception plus Tactile II using PNS and TENS stimulation simultaneously.
