## Supplemental File 4. for "Restoration of Natural Somatic Sensations to the Amputees: Finding the Right Combination of Neurostimulation Methods"

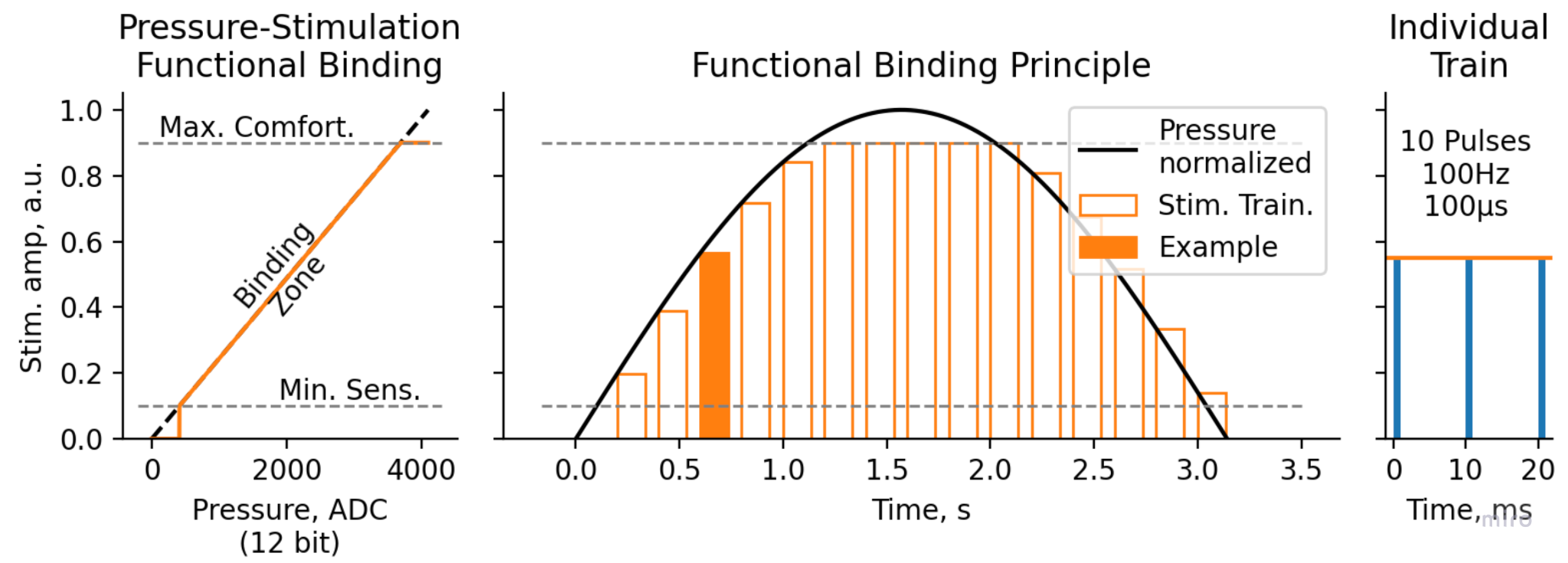

**Supplementary figure S4. Functional binding principle for Softness and Object size detection test.**

Pressure values, measured with 12-bit ADC ([0..4095] range), were then translated to electrostimulation amplitude linearly between Minimal Sensitive Stimulation threshold (1-2 Pts) and Maximal Comfortable Stimulation threshold (7-8 Pts). The stimulation were performed in trains of 10 pulses 100µs each, with pulse rate of 100Hz and train rate 4.6 Hz
